# The United States COVID-19 Forecast Hub dataset

**DOI:** 10.1101/2021.11.04.21265886

**Authors:** Estee Y Cramer, Yuxin Huang, Yijin Wang, Evan L Ray, Matthew Cornell, Johannes Bracher, Andrea Brennen, Alvaro J Castero Rivadeneira, Aaron Gerding, Katie House, Dasuni Jayawardena, Abdul H Kanji, Ayush Khandelwal, Khoa Le, Jarad Niemi, Ariane Stark, Apurv Shah, Nutcha Wattanchit, Martha W Zorn, Nicholas G Reich, on behalf of the US COVID-19 Forecast Hub Consortium

**Author notes:** corresponding author: Nicholas G Reich. These three authors contributed equally (Estee Y Cramer, Yuxin Huang, Yijin Wang). See group authorship list as appendix (on behalf of the US COVID-19 Forecast Hub Consortium). Disclaimer: The findings and conclusions in this report are those of the authors and do not necessarily represent the official position of the Centers for Disease Control and Prevention.

## Abstract

Academic researchers, government agencies, industry groups, and individuals have produced forecasts at an unprecedented scale during the COVID-19 pandemic. To leverage these forecasts, the United States Centers for Disease Control and Prevention (CDC) partnered with an academic research lab at the University of Massachusetts Amherst to create the US COVID-19 Forecast Hub. Launched in April 2020, the Forecast Hub is a dataset with point and probabilistic forecasts of incident hospitalizations, incident cases, incident deaths, and cumulative deaths due to COVID-19 at national, state, and county levels in the United States. Included forecasts represent a variety of modeling approaches, data sources, and assumptions regarding the spread of COVID-19. The goal of this dataset is to establish a standardized and comparable set of short-term forecasts from modeling teams. These data can be used to develop ensemble models, communicate forecasts to the public, create visualizations, compare models, and inform policies regarding COVID-19 mitigation. These open-source data are available via download from GitHub, through an online API, and through R packages.

## Background & Summary

To understand how the COVID-19 pandemic would progress in the United States, dozens of academic research groups, government agencies, industry groups, and individuals produced probabilistic forecasts for COVID-19 outcomes starting in March 2020.^1^ We have collected forecasts from over 82 modeling teams in a data repository, thus making forecasts easily accessible for COVID-19 response efforts and forecast evaluation. The data repository is called the United States (US) COVID-19 Forecast Hub (hereafter, Forecast Hub) and was created through a partnership between the US Centers for Disease Control and Prevention (CDC) and an academic research lab at the University of Massachusetts Amherst.

The Forecast Hub was launched in early April 2020 and contains real-time forecasts of reported COVID-19 cases, hospitalizations, and deaths. As of September 8, 2021, the Forecast Hub had collected nearly 65 million individual point or quantile predictions contained within over 4,600 submitted forecast files from over 100 unique models. The forecasts submitted each week reflected a variety of forecasting approaches, data sources, and underlying assumptions. There were no restrictions in place regarding the underlying information or code used to generate real-time forecasts. Each week, the latest forecasts were combined into an ensemble forecast (Figure 1) and all recent forecast data were updated on an official COVID-19 Forecasting page hosted by the US CDC.^2^ The ensemble models were also used in the weekly reports that are posted on the Forecast Hub website (https://covid19forecasthub.org/doc/reports/).

**Figure 1:**
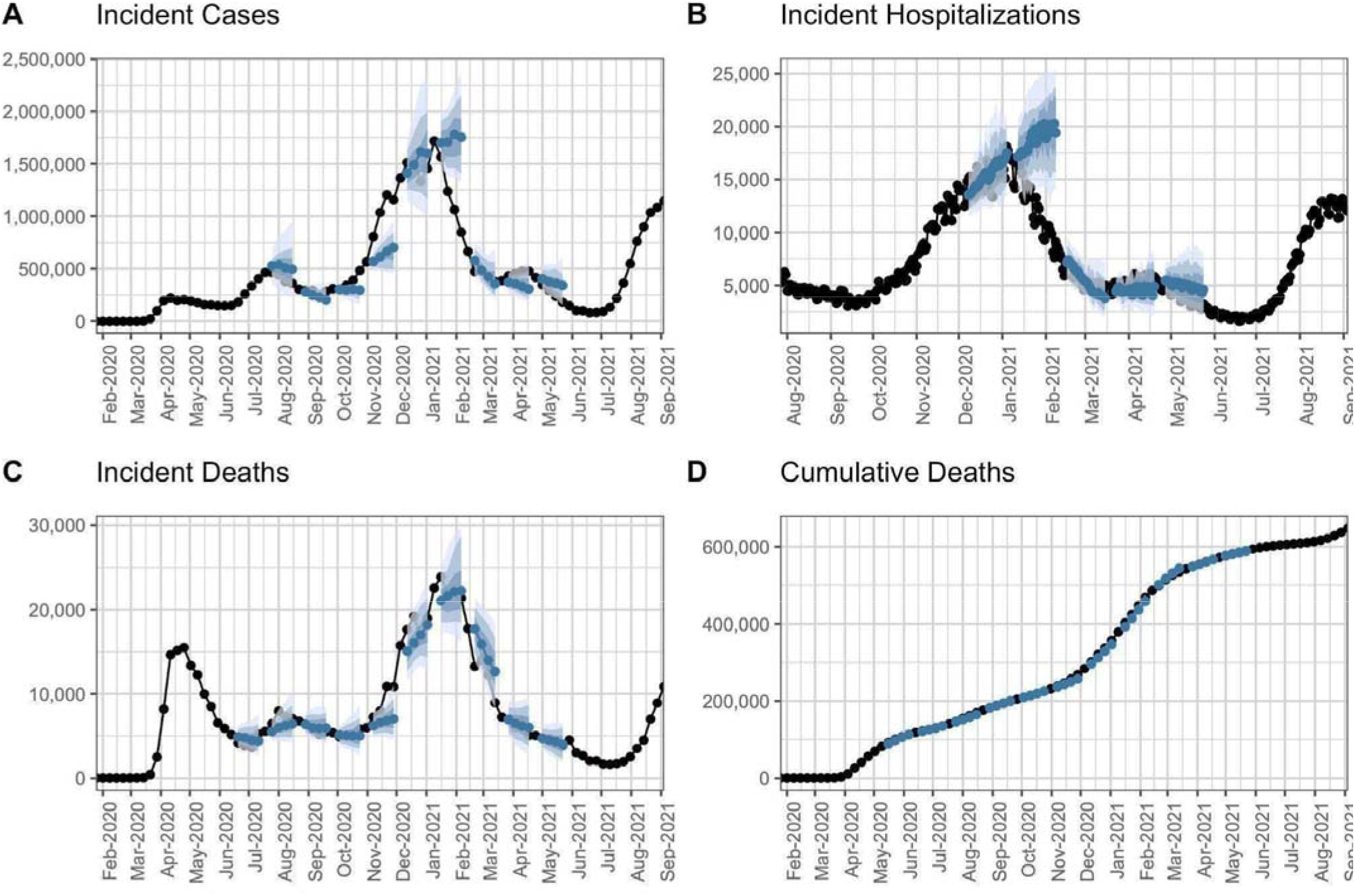
Time series of weekly incident deaths at the national level and forecasts from the COVID-19 Forecast Hub ensemble model for selected weeks in 2020 and 2021. Ensemble forecasts (blue) with 50%, 80% and 95% prediction intervals shown in shaded regions, and the ground-truth data (black) for incident cases (A), incident hospitalizations (B), incident deaths (C) and cumulative deaths (D). The truth data come from JHU CSSE (panels A, C, D) and HealthData.gov (panel B).

Forecasts are quantitative predictions about future observations. Forecasts differ from scenario-based projections, which examine feasible outcomes conditional on a variety of future assumptions. Because forecasts are unconditional estimates of future observations, they can be evaluated. An important feature of the Forecast Hub is that submitted forecasts are time-stamped so that the exact time at which a forecast was made public can be verified. In this way, the Forecast Hub serves as a public, independent registration system for these forecast model outputs. Data from the Forecast Hub have served as the basis for research articles for forecast evaluation^3^ and forecast combination.^4–6^ These studies can be used to determine how well models have performed at various points during the pandemic, which can, in turn, guide best practices for utilizing forecasts in practice and inform future forecasting efforts.^3^

Any modeling team was eligible to submit forecast data to the Forecast Hub, provided they submitted data in the correct format. Teams submitted predictions in a structured format to facilitate data validation, storage, and analysis. Teams also submitted a metadata file and license for their model’s data. Forecast data, ground truth data from the Johns Hopkins University Center for Systems Science and Engineering (JHU CSSE),^7^ New York Times (NYTimes),^8^ and USA Facts,^9^ as well as model metadata were stored in the public Forecast Hub GitHub repository.^10^

The forecasts were automatically synchronized with an online database called Zoltar via calls to a REpresentational State Transfer (REST) application programming interface (API)^11^ every six hours (Figure 2). Forecast data may be downloaded directly from GitHub, via the *covidHubUtils* R package,^12^ the zoltr R package^13^ or zoltpy python library.^14^

**Figure 2:**
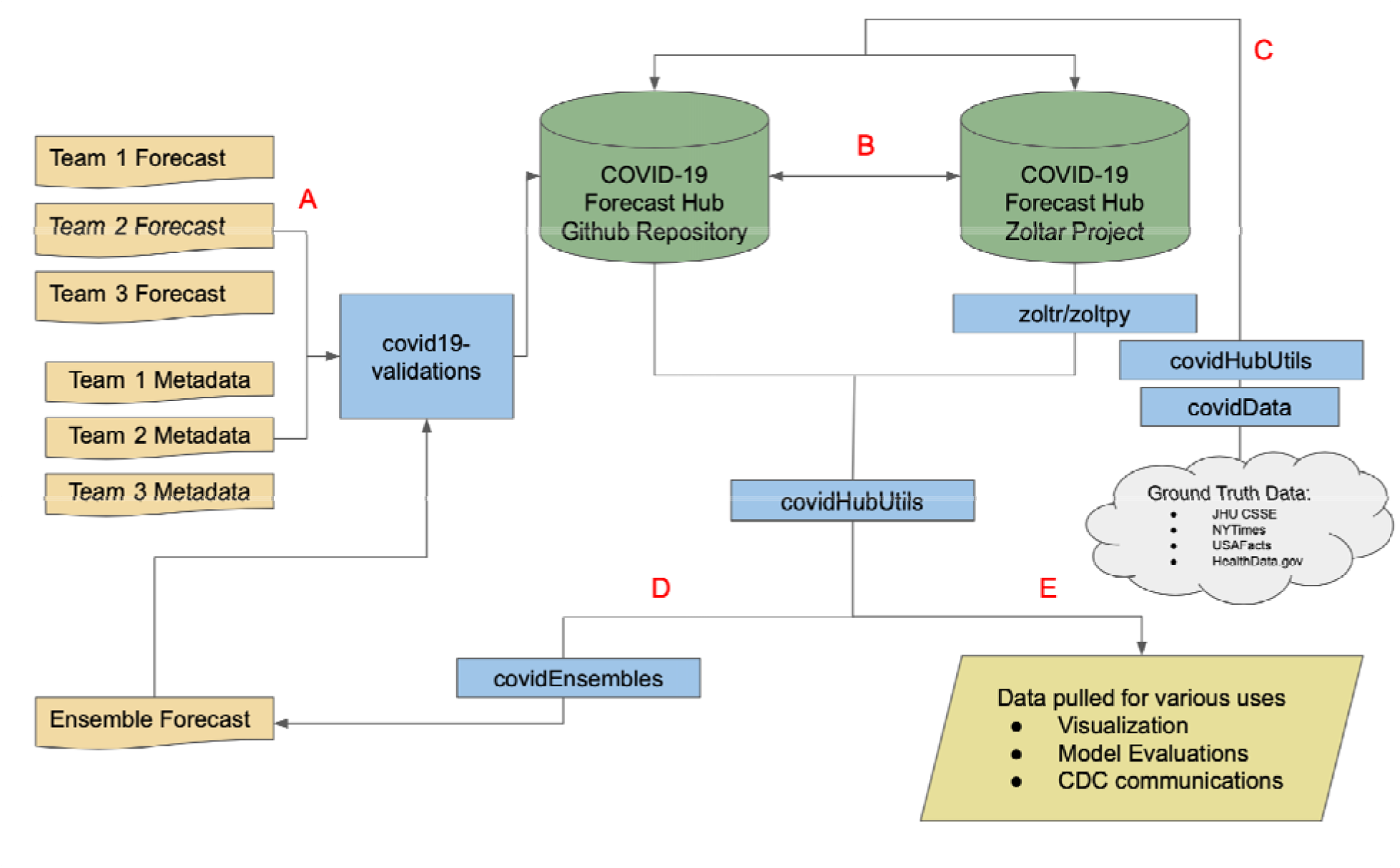
Schematic of the data storage and related infrastructure surrounding the COVID-19 Forecast Hub. (A) Forecasts are submitted to the COVID-19 Forecast Hub GitHub repository and undergo data format validation before being accepted into the system. (B) A continuous integration service ensures that the GitHub repository and PostgreSQL database stay in sync with essentially mirrored versions of the data. (C) Truth data for visualization, evaluation, and ensemble building are retrieved once per week using both the *covidHubUtils* and the *covidData* R packages. Truth data are stored in both repositories. (D) Once per week, an ensemble forecast submission is made using the *covidEnsembles* R package. It is submitted to the GitHub repository and undergoes the same validation as other submissions. (E) Using the *covidHubUtils* R package, forecast and truth data may be extracted from either the GitHub or PostgreSQL database in a standard format for tasks such as scoring or plotting.

This dataset of real-time forecasts created during the COVID-19 pandemic can provide insights into the shortcomings and successes of predictions and improve forecasting efforts in years to come. Though these data are restricted to forecasts for COVID-19 in the United States, the structure of this dataset has been used to create datasets of COVID-19 forecasts in the EU and the UK, and longer-term scenario projections in the US.^15–18^ The general structure of this data collection could be applied to additional diseases or forecasting outcome in the future.^11^

This large collaborative effort has provided data on short-term forecasts for over a year of forecasting efforts. These data were collected in real-time and therefore are not subject to retrospective biases. The data are also openly available to the public, thus fostering a transparent, open science approach to support public health efforts.

## Methods

### Data Acquisition

Beginning in April of 2020, the Reich Lab at the University of Massachusetts, Amherst, in partnership with the CDC, began collecting probabilistic forecasts of key COVID-19 outcomes in the United States (Table 1). The effort began by collecting forecasts of deaths and hospitalizations at the weekly and daily scale for the 50 US states, Washington DC, and 4 territories (Puerto Rico, US Virgin Islands, Guam, and the Northern Mariana Islands) as well as the aggregated US national level. In July 2020, the effort expanded to include forecasts of weekly incident cases at the county, state, and national levels. Forecasts may include a point prediction and/or quantiles of a predictive distribution.

**Table 1:** Forecast characteristics for all four outcomes. The table shows the temporal scale, spatial scale of locations, horizons stored, number of quantiles, and the dates of the earliest forecast, earliest standardized truth data, and the earliest ensemble build.

| Outcome | Scale | Locations |  |  | Horizons Stored | Number of quantiles for probabilistic forecasts | Earliest Forecast Date | First date of standardized truth data | Date of first ensemble forecast |
| --- | --- | --- | --- | --- | --- | --- | --- | --- | --- |
|  |  | County | State | National |  |  |  |  |  |
| Incident Cases | Weekly | X | X | X | 1 - 8 weeks | 7 | 2020-07-05 | 2020-03-15 | 2020-07-18 |
| Incident Hospitalizations | Daily |  | X | X | 1 - 130 days | 23 | 2020-03-27 | 2020-11-16 | 2020-12-05 |
| Incident Deaths | Daily |  | X | X | 1 - 130 days | 23 | 2020-03-15 | 2020-03-15 | NA |
| Incident Deaths | Weekly |  | X | X | 1-20 weeks | 23 | 2020-03-15 | 2020-03-15 | 2020-06-20 |
| Cumulative Deaths | Daily |  | X | X | 1 - 130 days | 23 | 2020-03-15 | 2020-03-15 | NA |
| Cumulative Deaths | Weekly |  | X | X | 1-20 weeks | 23 | 2020-03-15 | 2020-03-15 | 2020-04-13 |

Any team was eligible to submit data to the Forecast Hub. Upon initial submission of forecast data, teams were required to upload a metadata file that briefly described the methods used to create the forecasts and specified a license under which their forecast data were released. No model code was stored by the Forecast Hub.

During the first month of operation, members of the Forecast Hub team downloaded forecasts made available by teams publicly online, transformed these into the correct format, and pushed them into the Forecast Hub repository. Starting in May 2020, all teams were required to format and submit their own forecasts.

### Repository structure

The dataset is stored in two locations, and all data can be accessed through either source. The first is the COVID-19 Forecast Hub GitHub repository and the second is an online database, Zoltar, which can be accessed via a REST API.^11^ Details about data format and access are documented in the subsequent sections.

### Zoltar: data backend

The data can be accessed through the Zoltar forecast repository REST API. Through the API, subsets of submitted forecasts can be queried directly from a PostgreSQL database. This eliminates the need to access individual CSV files and facilitates access to versions of forecasts in cases when they are updated.

### Outcomes and locations

The Forecast Hub dataset stores forecasts for four different outcomes: incident hospitalizations, incident cases, incident deaths, and cumulative deaths (Table 1). Incident hospitalizations can be submitted for a horizons of 1 - 130 days in the future, incident cases can be submitted for 1 - 8 weeks in the future, and incident and cumulative deaths can be submitted for 1 - 20 weeks into the future. For all outcomes, forecasts can be submitted on a national and state level. Incident case forecasts were first introduced as a forecast outcome several months after the Hub started and have several key differences with other predicted outcomes. They are the only outcome for which the Hub accepts county-level forecasts in addition to the state and national level. Because there are over 3,000 counties in the US, this required some compromises on the scale of data collected for these forecasts in other ways. Specifically, case forecasts are required to have fewer quantiles (seven quantiles) compared to other outcomes which can have up to twenty-three quantiles. This gives a coarser representation of the forecast (see the section on Forecast format below).

Weekly targets follow the standard of epidemiological weeks (EW) used by the CDC, which defines a week to start on a Sunday and end on the following Saturday.^19^ Forecasts of cumulative deaths target the number of cumulative deaths reported by the Saturday ending a given week. Forecasts of weekly incident cases or deaths target the difference between reported cumulative cases or deaths on consecutive Saturdays. As an example of a forecast and the corresponding observation, forecasts submitted between Tuesday, October 6, 2020 (day 3 of EW41) and Monday, October 12, 2020 (day 2 of EW42) contained a “1 week ahead” forecast of incident deaths that corresponded to the change in cumulative reported deaths observed in EW42 (i.e., the difference between the cumulative reported deaths on Saturday, October 17, 2020, and Saturday, October 10, 2020), a “2 week ahead” forecast that corresponded to the change in cumulative reported deaths in week EW43. In this paper, we refer to the “forecast week” of a submitted forecast as the week corresponding to a “0-week ahead” horizon. In the example above, the forecast week would be EW41. Daily incident hospitalization horizons are for the number of reported hospitalizations a specified number of days after the forecast was generated.

### Forecast assumptions

Forecasters used a variety of assumptions to build models and generate predictions. Forecasting approaches include statistical or machine learning models, mechanistic models incorporating disease transmission dynamics, and combinations of multiple approaches.^3^ Teams have also included varying assumptions regarding future changes in policies and physical distancing measures, the transmissibility of COVID-19, vaccination rates, and the spread of new virus variants throughout the United States.

### Weekly submissions

A forecast submission consists of a single comma-separated value (CSV) file submitted via pull request to the GitHub repository. Forecast submissions are validated for technical accuracy and formatting (see exclusion criteriabelow) before being merged. To be included in the weekly ensemble model, teams were required to submit their forecast on Sunday or prior to a deadline on Monday. The majority of teams contributing to the dataset submitted forecasts to the Hub repository on Sunday or Monday, although some teams submitted at other times depending on their model production schedule.

### Model designation

Each model stored in the repository must have a classification of “primary”, “secondary” “other”. Each team must only have one “primary” model. Teams submitting multiple models with similar forecasting approaches can use the designations “secondary” or “other” for their models. Models with the designation “primary” are included in evaluations, the weekly ensemble, and the visualization. The “secondary” label is designed for models that have a substantive methodological difference than a team’s “primary” model. Models with the designation “secondary” are included only in the weekly ensemble and the visualization. The “other” label is designed for models that are small variations on a team’s “primary” model. Models with the designation “other” are not included in evaluations, the ensemble build, or the visualization.

### Ensemble and baseline forecasts

Several models have a special status, either as a baseline or as an ensemble that combines multiple models from the Hub to create a single forecast.

The COVIDhub-baseline model was created by the Hub in May 2020 as a benchmarking model. Its point forecast is the most recent observed value as of the forecast creation date with a probability distribution around that based on weekly differences in previous observations.^3^ The baseline model initially produced forecasts for case and death outcomes. Hospitalization baseline forecasts were added in September 2021.

The COVIDhub-ensemble model creates a combination of submitted forecasts to the Hub. Other work details the methods used for determining the appropriate combination approach.^4,5^ Starting in February 2021, GitHub tags were created to document the exact version of the repository used each week to create the COVIDhub-ensemble forecast. This creates an auditable trail in the repository so the correct version of the used forecasts could be recovered even in cases when some forecasts were subsequently updated.

Several other models also are combinations of some or all models submitted to the Forecast Hub. As of August 1, 2021, these models are COVIDhub-trained_ensemble, FDANIHASU-Sweight, JHUAPL-SLPHospEns, and KITmetricslab-select_ensemble. These models are flagged in the metadata using the Boolean metadata field, “ensemble_of_hub_models”.

### Exclusion criteria

No forecasts were excluded from the dataset due to the forecast values or the background experience of the forecasters. Forecast files were only rejected if they did not meet the automatic formatting criteria implemented through automatic GitHub checks.^20^ These included checks to ensure that, among other criteria:

- A forecast file is submitted no more than 2 days after it has been created (to ensure forecasts submitted were truly prospective). The creation date is based on the date in the filename created by the submitting team.
- The forecast dates in the content of the file are in the format YYYY-MM-DD and match the creation date.
- Quantile forecasts do not contain any quantiles at probability levels other than those required (see Forecast Format section below).

### Updates to files

To ensure that forecasting is done in real-time, all forecasts are required to be submitted to the Hub within 2 days of the forecast date, which is listed in a column within each forecast file. Though occasional late submissions were accepted up through January of 2021, the policy was updated to not accept late forecasts due to missed deadlines, updated modeling methods, or other reasons.

Exceptions to this policy were made if there were programing or data errors that affected the forecasts in the original submission or if a new team joined. If there was an error, teams were required to submit a comment with their updated submission affirming that there was a bug and that the forecast was only produced using data that were available at the time of the original submission. In the case of updates to forecast data, both the old and updated versions of the forecasts can be accessed either through the GitHub commit history or through time-stamped queries of the forecasts in the Zoltar database. Note that an updated forecast can include “retracting” a particular set of predictions in the case when an initial forecast was not able to be updated. When new teams join the Hub, they can submit late forecasts if they can provide publicly available evidence that the forecasts were made in real-time (e.g. GitHub commit history).

### Ground truth data

Data from the JHU CSSE dataset^21^ are used as the ground truth data for reported cases and deaths. Data from the HealthData.gov system for state-level hospitalizations are used for the hospitalization outcome. JHU CSSE obtained counts of cases and deaths by collecting and aggregating reports from state and local health departments. HealthData.gov contains reports of hospitalizations assembled by the U.S. Department of Health and Human Services. Teams were encouraged to use these sources to build models. Although hospitalization forecasts were collected starting in March 2020, the hospitalizations data from HealthData.gov were only available later, and we started encouraging teams to target these data in November 2020. Some teams used alternate data sources including the NYTimes, USAFacts, US Census data, and other signals.^3^ Versions of truth data from JHU CSSE, USAFacts, and the NYTimes are stored in the GitHub repository.

Previous reports of ground truth data for past time points were occasionally updated as new records became available, definitions of reportable cases, deaths, or hospitalizations changed, or errors in data collection were identified and corrected. These revisions to the data are sometimes quite substantial, and for some purposes such as retrospective ensemble construction, it is necessary to use the data that would have been available in real-time. The historically versioned data can be accessed either through GitHub commit records, data versions released on HealthData.gov, or third-party tools such as the covidcast API provided by the Delphi group at Carnegie Mellon University or the *covidData* R package.^22^

## Data Records

### Summary of forecast data collected

In the initial weeks of submission, there were fewer than 10 models providing forecasts. As the pandemic spread, the number of teams submitting forecasts increased to 82; as of July 2021, 82 primary, 4 secondary models and 15 models with the designation “other” had been submitted to the Forecast Hub. In the first six months of 2021, a median of 35.5 teams (range: 30 to 38) contributed incident case forecasts (Fig 3a), a median of 12 teams (range: 9 to 14) contributed incident hospitalizations (Fig 3b), a median of 43 teams (range 37 to 49) contributed incident death forecasts (Fig 3c), and a median of 44 teams (range 34 to 46) contributed cumulative death forecasts (Fig 3d). As of September 8 2021, the dataset contained 4,602 forecast files with 64,902,239 point or quantile predictions for unique combinations of targets and locations.

**Figure 3:**
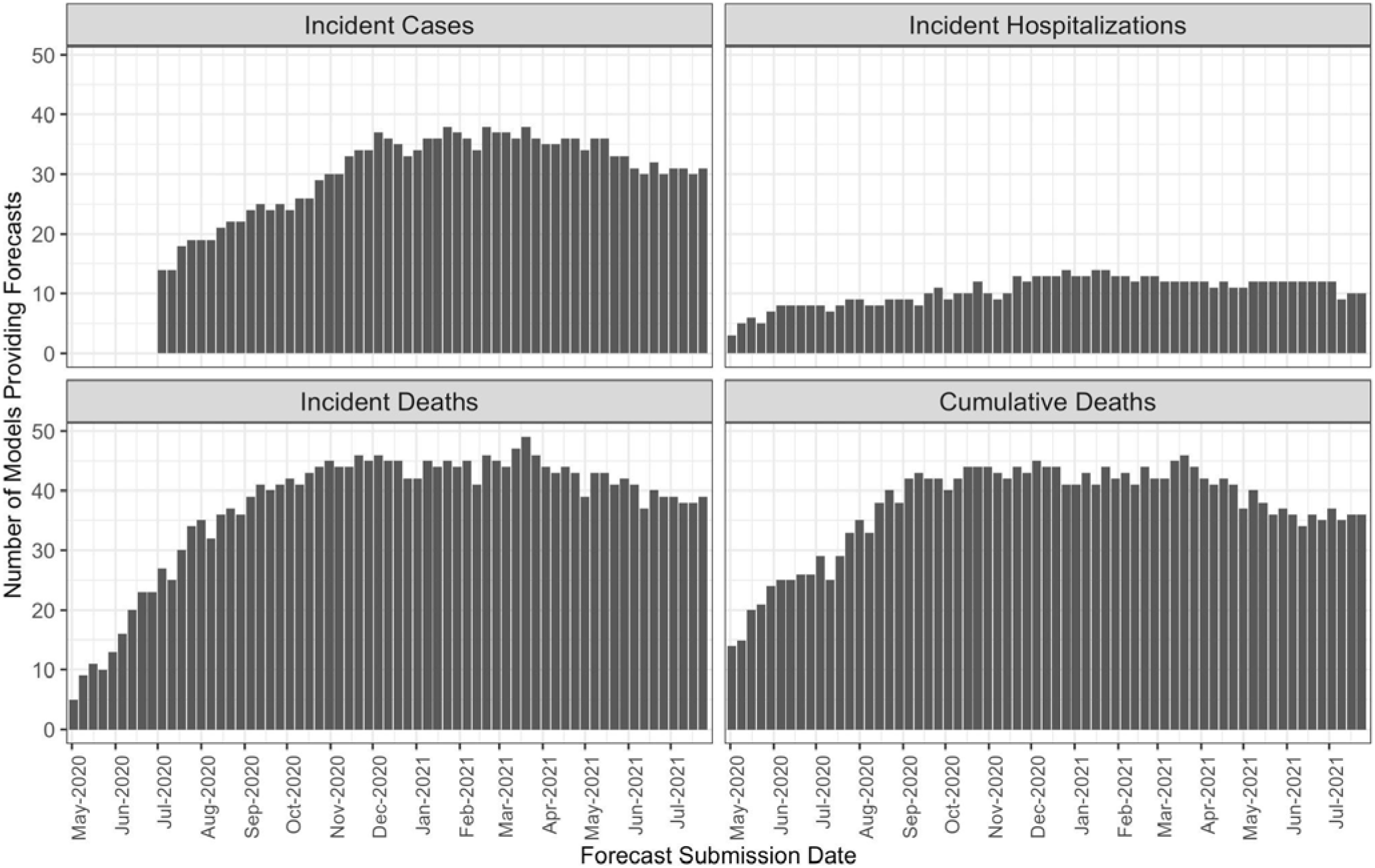
Number of primary forecasts submitted for each outcome per week from April 27th 2020 through July 31st 2021. In the initial weeks of submission, fewer than 10 models provided forecasts. Over time, the number of teams submitting forecasts for each forecasted outcome increased into early 2021 and then saw a small decline through the summer of 2021.

### GitHub repository data structure

Forecasts in the GitHub repository are available in subfolders organized by model. Folders are named with a team name and model name, and each folder includes a metadata file and forecast files. Forecast CSV files are named using the format “<YYYY-MM-DD>-<team abbreviation>-<model abbreviation>.csv”. In these files, each row contains data for a single outcome, location, horizon, and point or quantile prediction as described above.

The metadata file for each team, named using the format “metadata-<team abbreviation>-<model abbreviation>.txt”, contains relevant information about the team and the model that the team is using to generate forecasts.

### Forecast format

Forecasts were required to be submitted in the format of point predictions and/or quantile predictions. Point predictions represented single “best” predictions with no uncertainty, typically representing a mean or median prediction from the model. Quantile predictions are an efficient format for storing predictive distributions of a wide range of outcomes.

Quantile representations of predictive distributions lend themselves to natural computations of, for example, pinball loss or a weighted interval score, both strictly proper scoring rules that can be used to evaluate forecasts^23^ However, they do not capture the structure of the tails of the predictive distribution beyond the reported quantiles. Also, the quantile format does not preserve any information on correlation structures between different outcomes.

### Variable descriptions

The forecast data in this dataset are stored in seven columns:

1. **forecast_date** - the date the forecast was made in the format YYYY-MM-DD.
2. **target** - a character string giving the number of days/weeks ahead that are being forecasted (horizon) and the outcome. Horizons must be one of the following:
  a. “N wk ahead cum death” where N is a number between 1 and 20
  b. “N wk ahead inc death” where N is a number between 1 and 20
  c. “N wk ahead inc case” where N is a number between 1 and 8
  d. “N day ahead inc hosp” where N is a number between 0 and 130
3. **target_end_date** - a character string representing the date for the forecast target in the format YYYY-MM-DD. For “k day-ahead” targets, target_end_date will be k days after forecast_date. For “k week ahead” targets, target_end_date will be the Saturday at the end of the specified epidemic week, as described above.
4. **location** - character string of Federal Information Processing Standard Publication (FIPS) codes identifying U.S. states, counties, territories, and districts as well as “US” for national forecasts. The values for the FIPS codes are available in a CSV file in the repository and as a data object in the *covidHubUtils* R package for convenience.
5. **type** - character value of “point” or “quantile” indicating whether the row corresponds to a point forecast or a quantile forecast.
6. **quantile** - the probability level for a quantile forecast. For death and hospitalization forecasts, forecasters can submit quantiles at 23 probability levels: 0.01, 0.025, 0.05, 0.10, 0.15, …, 0.95, 0.975, 0.99. For cases, teams can submit up to 7 quantiles at levels .025, 0.100, 0.250, 0.5, 0.750, 0.900 and 0.975. If the forecast “type” is equal to “point”, the value in the quantile column is equal to “NA”.
7. **value** - non-negative numbers indicating the “point” or “quantile” prediction for the row. For a “point” prediction, value is simply the value of that point prediction for the target and location associated with that row. For a “quantile” prediction, the model predicts that the eventual observation will be less than this value with the probability given by the quantile probability level.

### Metadata format

Each team documents their model information in a metadata file which is required along with the first forecast submission. Each team is asked to record their model’s design and assumptions, the model contributors, the team’s website, information regarding the team’s data sources, and a brief model description. Teams may update their metadata file periodically to keep track of minor changes to a model.

### Variable descriptions

A standard metadata file should be a YAML file with the following required fields in a specific order:

1. **team_name** - the name of the team (less than 50 characters).
2. **model_name** - the name of the model (less than 50 characters).
3. **model_abbr** - an abbreviated and uniquely identified name for the model that is less than 30 alphanumeric characters. The model abbreviation must be in the format of ‘[team_abbr]-[model_abbr]’ where each of the ‘[team_abbr]’ and ‘[model_abbr]’ are text strings that are each less than 15 alphanumeric characters that do not include a hyphen or whitespace.
4. **model_contributors** - a list of all individuals involved in the forecasting effort, affiliations, and email addresses. At least one contributor needs to have a valid email address. The syntax of this field should be name1 (affiliation1) <user@address>, name2 (affiliation2) <user2@address2>
5. **website_url\*** - a URL to a website that has additional data about the model. We encourage teams to submit the most user-friendly version of the model, e.g. a dashboard, or similar, that displays the model forecasts. If there is an additional data repository where forecasts and other model code are stored, this can be included in the methods section. If only a more technical site, e.g. GitHub repo, exists that link should be included here.
6. **license** - one of the acceptable license types in the Forecast Hub. We encourage teams to submit as a “cc-by-4.0” to allow the broadest possible use, including private vaccine production (which would be excluded by the “cc-by-nc-4.0” license). If the value is “LICENSE.txt”, then a LICENSE.txt file must exist within the model folder and provide a license.
7. **team_model_designation** - upon initial submission this field should be one of “primary”, “secondary” or “other”.
8. **methods** - a brief description of the forecasting methodology that is less than 200 Characters.
9. **ensemble_of_hub_models** - a Boolean value (‘true’ or ‘false’) that indicates whether a model combines multiple hub models into an ensemble.

*previously named **model_output**

Teams are also encouraged to add model information with optional fields described in Supplement 1.

## Technical Validation

Two similar but distinct validation processes were used to validate data on the GitHub repository and on Zoltar.

### GitHub repository

Validations were set up using GitHub Actions to manage continuous integration and automated data checking.^20^ Teams submitted their metadata files and forecasts through pull requests on GitHub. Each time a new pull request was submitted, a validation script ran on all new or updated files in the pull request to test for their validity. Separate checks ran on metadata file changes and forecast data file changes.

The metadata file for each team was required to be in valid YAML format, and a set of specific checks were required before a new metadata file could be merged into the repository. Checks included ensuring that the proposed team and model names do not conflict with existing names, that a valid license for data reuse is specified, and that a valid model designation was present. A list of specific validations for metadata may be found in Supplement 2.

New or changed forecast data files for each team were required to pass a series of checks for data formatting and validity. These checks also ensured that the forecast data files did not meet any of the exclusion criteria (see the Methods section for specific rules). Furthermore, a list of specific validations for forecast data files is provided in Supplement 2.

### Zoltar

When a new forecast file is uploaded to Zoltar, unit tests are run on the file to make sure that forecast elements contain valid structure. (For a detailed specification of the structure of forecast elements, see https://docs.zoltardata.com/validation/.) If a forecast file does not pass all unit tests, the upload will fail and the forecast file will not be added to the database; only when all tests pass will the new forecast be added to Zoltar. The validations in place on GitHub ensure that only valid forecasts will be uploaded to Zoltar.

### Observed data

Raw observed data from multiple sources including JHU, NYTimes, USAFacts, and Healthdata.gov is downloaded and reformatted using the scripts in the R packages *covidHubUtils* (https://github.com/reichlab/covidHubUtils) and *covidData* (https://github.com/reichlab/covidData. This data generating process is automated by GitHub Actions every week and the results (called “truth data”) are directly uploaded to the Forecast Hub repository and Zoltar. In specific, case and death raw observed data are aggregated to a weekly level and all three outcomes (cases, deaths, and hospitalization) are reformatted for use within the Hub.

## Usage Notes

We have developed the *covidHubUtils* R package (https://github.com/reichlab/covidHubUtils) to facilitate bulk retrieval of forecasts for analysis and evaluation. Examples of how to use the *covidHubUtils* package and its functions can be found at https://reichlab.io/covidHubUtils/. The package supports loading forecasts from a local clone of the GitHub repository or by querying data from Zoltar. The package supports common actions for working with the data, such as loading in specific subsets of forecasts, plotting forecasts, scoring forecasts, retrieving ground truth data, and many other utility functions to simplify working with the data.

### Communicating results from the COVID-19 Forecast Hub

Communication of probabilistic forecasts to the public is challenging, ^24,25^ and the best practices regarding the communication of outbreaks are still developing.^26^ Starting in April 2020, the CDC published weekly summaries of these forecasts on their public website^27^, and these forecasts were occasionally used in public briefings by the CDC director.^28^ Additional examples of the communication of Forecast Hub data can be viewed through weekly reports generated by the Hub team for dissemination to the general public, including state and local departments of health.(https://covid19forecasthub.org/doc/reports/)

## Data Availability

All data produced are available online at https://github.com/reichlab/covid19-forecast-hub

## Acknowledgements

This work has been supported in part by the US Centers for Disease Control and Prevention (1U01IP001122) and the National Institutes of General Medical Sciences (R35GM119582). The content is solely the responsibility of the authors and does not necessarily represent the official views of CDC, FDA, NIGMS or the National Institutes of Health. Johannes Bracher was supported by the Helmholtz Foundation via the SIMCARD Information & Data Science Pilot Project. Tilmann Gneiting gratefully acknowledges support by the Klaus Tschira Foundation.

## Funding

For teams that reported receiving funding for their work, we report the sources and disclosures below.

*<u>AIpert-pwllnod</u>*:Natural Sciences and Engineering Research Council of Canada

*<u>Caltech-CS156</u>*: Gary Clinard Innovation Fund

*<u>CEID-Walk</u>*: University of Georgia

*<u>CMU-TimeSeries</u>*: CDC Center of Excellence, gifts from Google and Facebook

*<u>COVIDhub</u>*:This work has been supported by the US Centers for Disease Control and Prevention (1U01IP001122) and the National Institutes of General Medical Sciences (R35GM119582). The content is solely the responsibility of the authors and does not necessarily represent the official views of CDC, NIGMS or the National Institutes of Health. Johannes Bracher was supported by the Helmholtz Foundation via the SIMCARD Information & Data Science Pilot Project. Tilmann Gneiting gratefully acknowledges support by the Klaus Tschira Foundation.

*<u>CU-select:</u>* NSF DMS-2027369 and a gift from the Morris-Singer Foundation.

*<u>DDS-NBDS</u>*:NSF III-1812699

*<u>epiforecasts-ensemble1</u>*: Wellcome Trust (210758/Z/18/Z)

*<u>FDANIHASU</u>*: supported by the Intramural Research Program of the NIH/NIDDK

*<u>GT_CHHS-COVID19</u>*: William W. George Endowment, Virginia C. and Joseph C. Mello Endowment, NSF DGE-1650044, NSF MRI 1828187, research cyberinfrastructure resources and services provided by the Partnership for an Advanced Computing Environment (PACE) at Georgia Tech, and the following benefactors at Georgia Tech: Andrea Laliberte, Joseph C. Mello, Richard “Rick” E. & Charlene Zalesky, and Claudia & Paul Raines, CDC MInD-Healthcare U01CK000531-Supplement.

*<u>IHME</u>*: This work was supported by the Bill & Melinda Gates Foundation, as well as funding from the state of Washington and the National Science Foundation (award no. FAIN: 2031096)

<u>Imperial-ensemble1:</u>SB acknowledges funding from the Wellcome Trust (219415).

*<u>Institute of Business Forecasting</u>* : IBF

*<u>IowaStateLW-STEM</u>*: NSF DMS-1916204, Iowa State University Plant Sciences Institute Scholars Program, NSF DMS-1934884, Laurence H. Baker Center for Bioinformatics and Biological Statistics.

*<u>IUPUI CIS :</u>* NSF

*<u>JHU_CSSE-DECOM</u>*: JHU CSSE: National Science Foundation (NSF) RAPID “Real-time Forecasting of COVID-19 risk in the USA”. 2021-2022. Award ID: 2108526. National Science Foundation (NSF) RAPID “Development of an interactive web-based dashboard to track COVID-19 in real-time”. 2020. Award ID: 2028604

*<u>JHU_IDD-CovidSP</u>*: State of California, US Dept of Health and Human Services, US Dept of Homeland Security, Johns Hopkins Health System, Office of the Dean at Johns Hopkins Bloomberg School of Public Health, Johns Hopkins University Modeling and Policy Hub, Centers for Disease Control and Prevention (5U01CK000538-03), University of Utah Immunology, Inflammation, & Infectious Disease Initiative (26798 Seed Grant).

*<u>JHU_UNC_GAS-StatMechPool</u>*: NIH NIGMS: R01GM140564

*<u>JHUAPL-Bucky</u>*: US Dept of Health and Human Services

*<u>KITmetricslab-select_ensemble</u>*: Daniel Wolffram gratefully acknowledges support by the Klaus Tschira Foundation.

*<u>LANL-GrowthRate</u>*: LANL LDRD 20200700ER

*<u>MIT-Cassandra</u>*: MIT Quest for Intelligence

*<u>MOBS-GLEAM_COVID</u>*: COVID Supplement CDC-HHS-6U01IP001137-01; CA

NU38OT000297 from the Council of State and Territorial Epidemiologists (CSTE)

*<u>NotreDame-FRED</u>*: NSF RAPID DEB 2027718

*<u>NotreDame-mobility</u>*: NSF RAPID DEB 2027718

*<u>PSI-DRAFT</u>*: NSF RAPID Grant # 2031536

*<u>QJHong-Encounter</u>*: NSF DMR-2001411 and DMR-1835939

*<u>SDSC_ISG-TrendModel</u>*: The development of the dashboard was partly funded by the Fondation Privée des Hôpitaux Universitaires de Genève

*<u>UA-EpiCovDA</u>*: NSF RAPID Grant # 2028401

UChicagoCHATTOPADHYAY-UnIT: Defense Advanced Research Projects Agency (DARPA) #HR00111890043/P00004 (I. Chattopadhyay, University of Chicago).

*<u>UCSB-ACTS</u>*: NSF RAPID IIS 2029626

*<u>UCSD_NEU-DeepGLEAM</u>*: Google Faculty Award, W31P4Q-21-C-0014

*<u>UMass-MechBayes</u>*: NIGMS #R35GM119582, NSF #1749854, NIGMS #R35GM119582

*<u>UMich-RidgeTfReg</u>*: This project is funded by the University of Michigan Physics Department and the University of Michigan Office of Research.

*<u>UVA-Ensemble</u>*: National Institutes of Health (NIH) Grant 1R01GM109718, NSF BIG DATA Grant IIS-1633028, NSF Grant No.: OAC-1916805, NSF Expeditions in Computing Grant CCF-1918656, CCF-1917819, NSF RAPID CNS-2028004, NSF RAPID OAC-2027541, US Centers for Disease Control and Prevention 75D30119C05935, a grant from Google, University of Virginia Strategic Investment Fund award number SIF160, Defense Threat Reduction Agency (DTRA) under Contract No. HDTRA1-19-D-0007, and Virginia Dept of Health Grant VDH-21-501-0141

*<u>Wadnwani_AI-BayesOpt</u>*: This study is made possible by the generous support of the American People through the United States Agency for International Development (USAID). The work described in this article was implemented under the TRACETB Project, managed by WIAI under the terms of Cooperative Agreement Number 72038620CA00006. The contents of this manuscript are the sole responsibility of the authors and do not necessarily reflect the views of USAID or the United States Government.

*<u>WalmartLabsML-LogForecasting</u>*: Team acknowledges Walmart to support this study

## Author Consortium

Estee Y Cramer^1^, Yuxin Huang^1^, Yijin Wang^1^, Evan L Ray^1^, Matthew Cornell^1^, Johannes Bracher^2,3^, Andrea Brennen^4^, Alvaro J Castro Rivadeneira^1^, Aaron Gerding^1^, Katie House^1^, Dasuni Jayawardena^1^, Abdul H Kanji^1^, Ayush Khandelwal^1^, Khoa Le^1^, Jarad Niemi^5^, Ariane Stark^1^, Apurv Shah^1^, Nutcha Wattanachit^1^, Martha W Zorn^1^, Tilmann Gneiting^2^, Anja Mühlemann^6^, Youyang Gu^7^, Yixian Chen^8^, Krishna Chintanippu^8^, Viresh Jivane^8^, Ankita Khurana^8^, Ajay Kumar^8^, Anshul Lakhani^8^, Prakhar Mehrotra^8^, Sujitha Pasumarty^8^, Monika Shrivastav^8^, Jialu You^8^, Nayana Bannur^9^, Ayush Deva^9^, Sansiddh Jain^9^, Mihir Kulkarni^9^, Srujana Merugu^9^, Alpan Raval^9^, Siddhant Shingi^9^, Avtansh Tiwari^9^, Jerome White^9^, Aniruddha Adiga^10^, Benjamin Hurt^10^, Bryan Lewis^10^, Madhav Marathe^10^, Akhil Sai Peddireddy^10^, Przemyslaw Porebski^10^, Srinivasan Venkatramanan^10^, Lijing Wang^10^, Maytal Dahan^11^, Spencer Fox^12^, Kelly Gaither^11^, Michael Lachmann^13^, Lauren Ancel Meyers^12^, James G Scott^12^, Mauricio Tec^12^, Spencer Woody^12^, Ajitesh Srivastava^14^, Tianjian Xu^14^, Jeffrey C Cegan^15^, Ian D Dettwiller^15^, William P England^15^, Matthew W Farthing^15^, Glover E George^15^, Robert H Hunter^15^, Brandon Lafferty^15^, Igor Linkov^15^, Michael L Mayo^15^, Matthew D Parno^15^, Michael A Rowland^15^, Benjamin D Trump^15^, Samuel Chen^16^, Stephen V Faraone^16^, Jonathan Hess^16^, Christopher P Morley^16^, Asif Salekin^17^, Dongliang Wang^16^, Yanli Zhang-James^16^, Thomas M Baer^18^, Sabrina M Corsetti^19^, Marisa C Eisenberg^19^, Karl Falb^19^, Yitao Huang^19^, Emily T Martin^19^, Ella McCauley^19^, Robert L Myers^19^, Tom Schwarz^19^, Graham Casey Gibson^1^, Daniel Sheldon^1^, Liyao Gao^20^, Yian Ma^21^, Dongxia Wu^21^, Rose Yu^22,21^, Xiaoyong Jin^23^, Yu-Xiang Wang^23^, Xifeng Yan^23^, YangQuan Chen^24^, Lihong Guo^25^, Yanting Zhao^26^, Jinghui Chen^27^, Quanquan Gu^27^, Lingxiao Wang^27^, Pan Xu^27^, Weitong Zhang^27^, Difan Zou^27^, Ishanu Chattopadhyay^28^, Yi Huang^28^, Guoqing Lu^29^, Ruth Pfeiffer^30^, Timothy Sumner^31^, Liqiang Wang^31^, Dongdong Wang^31^, Shunpu Zhang^31^, Zihang Zou^31^, Hannah Biegel^32^, Joceline Lega^32^, Fazle Hussain^33^, Zeina Khan^33^, Frank Van Bussel^33^, Steve McConnell^34^, Stephanie L Guertin^35^, Christopher Hulme-Lowe^35^, VP Nagraj^35^, Stephen D Turner^35^, Benjamín Bejar^36^, Christine Choirat^36^, Antoine Flahault^37^, Ekaterina Krymova^36^, Gavin Lee^36^, Elisa Manetti^37^, Kristen Namigai^37^, Guillaume Obozinski^36^, Tao Sun^36^, Dorina Thanou^38^, Xuegang Ban^20^, Yunfeng Shi^39^, Robert Walraven^7^, Qi-Jun Hong^40,41^, Axel van de Walle^41^, Michal Ben-Nun^42^, Steven Riley^43^, Pete Riley^42^, James A Turtle^42^, Duy Cao^44^, Joseph Galasso^44^, Jae H Cho^7^, Areum Jo^7^, David DesRoches^45^, Pedro Forli^45^, Bruce Hamory^45^, Ugur Koyluoglu^45^, Christina Kyriakides^45^, Helen Leis^45^, John Milliken^45^, Michael Moloney^45^, James Morgan^45^, Ninad Nirgudkar^45^, Gokce Ozcan^45^, Noah Piwonka^45^, Matt Ravi^45^, Chris Schrader^45^, Elizabeth Shakhnovich^45^, Daniel Siegel^45^, Ryan Spatz^45^, Chris Stiefeling^45^, Barrie Wilkinson^45^, Alexander Wong^45^, Sean Cavany^46^, Guido España^46^, Sean Moore^46^, Rachel Oidtman^28,46^, Alex Perkins^46^, Andrea Kraus^47^, David Kraus^47^, Jiang Bian^48^, Wei Cao^48^, Zhifeng Gao^48^, Juan Lavista Ferres^48^, Chaozhuo Li^48^, Tie-Yan Liu^48^, Xing Xie^48^, Shun Zhang^48^, Shun Zheng^48^, Matteo Chinazzi^49^, Alessandro Vespignani^50,49^, Xinyue Xiong^49^, Jessica T Davis^49^, Kunpeng Mu^49^, Ana Pastore y Piontti^49^, Jackie Baek^51^, Vivek Farias^52^, Andreea Georgescu^51^, Retsef Levi^52^, Deeksha Sinha^51^, Joshua Wilde^51^, Andrew Zheng^51^, Amine Bennouna^51^, David Nze Ndong^52^, Georgia Perakis^53^, Divya Singhvi^54^, Ioannis Spantidakis^51^, Leann Thayaparan^51^, Asterios Tsiourvas^51^, Shane Weisberg^51^, Ali Jadbabaie^55^, Arnab Sarker^55^, Devavrat Shah^55^, Leo A Celi^56^, Nicolas D Penna^56^, Saketh Sundar^57^, Russ Wolfinger^58^, Lauren Castro^59^, Geoffrey Fairchild^59^, Isaac Michaud^59^, Dave Osthus^59^, Daniel Wolffram^2,3^, Dean Karlen^60,61^, Mark J Panaggio^62^, Matt Kinsey^62^, Luke C. Mullany^62^, Kaitlin Rainwater-Lovett^62^, Lauren Shin^62^, Katharine Tallaksen^62^, Shelby Wilson^62^, Michael Brenner^63,64^, Marc Coram^64^, Jessie K Edwards^65^, Keya Joshi^66^, Ellen Klein^64^, Juan Dent Hulse^67^, Kyra H Grantz^67^, Alison L Hill^68^, Joshua Kaminsky^67^, Kathryn Kaminsky^7^, Lindsay T Keegan^69^, Stephen A Lauer^67^, Elizabeth C Lee^67^, Joseph C Lemaitre^70^, Justin Lessler^71^, Hannah R Meredith^67^, Javier Perez-Saez^67^, Sam Shah^7^, Claire P Smith^67^, Shaun A Truelove^67^, Josh Wills^7^, Lauren Gardner^68^, Maximilian Marshall^68^, Kristen Nixon^68^, John C. Burant^7^, Wen-Hao Chiang^72^, George Mohler^72^, Junyi Gao^73^, Lucas Glass^74^, Cheng Qian^74^, Justin Romberg^75^, Rakshith Sharma^74^, Jeffrey Spaeder^76^, Jimeng Sun^73^, Cao Xiao^77^, Lei Gao^6^, Zhiling Gu^6^, Myungjin Kim^6^, Xinyi Li^78^, Guannan Wang^79^, Lily Wang^6^, Yueying Wang^6^, Shan Yu^10^, Chaman Jain^80^, Sangeeta Bhatia^81^, Pierre Nouvellet^81,82^, Ryan Barber^20^, Emmanuela Gaikedu^20^, Simon Hay^20^, Steve Lim^20^, Chris Murray^20^, David Pigott^20^, Robert C Reiner^20^, Prasith Baccam^83^, Heidi L Gurung^83^, Steven A Stage^83^, Bradley T Suchoski^83^, Chung-Yan Fong^84^, Dit-Yan Yeung^84^, Bijaya Adhikari^85^, Jiaming Cui^75^, B. Aditya Prakash^75^, Alexander Rodríguez^75^, Anika Tabassum^75,86^, Jiajia Xie^75^, John Asplund^87^, Arden Baxter^88^, Pinar Keskinocak^88^, Buse Eylul Oruc^88^, Nicoleta Serban^88^, Sercan O Arik^89^, Mike Dusenberry^89^, Arkady Epshteyn^89^, Elli Kanal^89^, Long T Le^89^, Chun-Liang Li^89^, Tomas Pfister^89^, Rajarishi Sinha^89^, Thomas Tsai^90^, Nate Yoder^89^, Jinsung Yoon^89^, Leyou Zhang^89^, Daniel Wilson^91^, Artur A Belov^92^, Carson C Chow^93^, Richard C Gerkin^40^, Osman N Yogurtcu^92^, Mark Ibrahim^94^, Timothee Lacroix^94^, Matthew Le^94^, Jason Liao^95^, Maximilian Nickel^94^, Levent Sagun^94^, Sam Abbott^96^, Nikos I Bosse^96^, Sebastian Funk^96^, Joel Hellewell^96^, Sophie R Meakin^96^, Katharine Sherratt^96^, Rahi Kalantari^97^, Mingyuan Zhou^97^, Sen Pei^98^, Jeffrey Shaman^98^, Teresa K Yamana^98^, Omar Skali Lami^51^, Dimitris Bertsimas^52^, Michael L Li^51^, Saksham Soni^51^, Hamza Tazi Bouardi^51^, Madeline Adee^99^, Turgay Ayer^100,88^, Jagpreet Chhatwal^101^, Ozden O Dalgic^102^, Mary A Ladd^99^, Benjamin P Linas^103^, Peter Mueller^99^, Jade Xiao^88^, Qinxia Wang^98^, Yuanjia Wang^98^, Shanghong Xie^98^, Donglin Zeng^104^, Jacob Bien^14^, Logan Brooks^105^, Alden Green^105^, Addison J Hu^105^, Maria Jahja^105^, Daniel McDonald^106^, Balasubramanian Narasimhan^107^, Collin Politsch^105^, Samyak Rajanala^107^, Aaron Rumack^105^, Noah Simon^20^, Ryan J Tibshirani^105^, Rob Tibshirani^107^, Valerie Ventura^105^, Larry Wasserman^105^, John M Drake^108^, Eamon B O’Dea^108^, Yaser Abu-Mostafa^109^, Rahil Bathwal^109^, Nicholas A Chang^109^, Pavan Chitta^109^, Anne Erickson^109^, Sumit Goel^109^, Jethin Gowda^109^, Qixuan Jin^109^, HyeongChan Jo^109^, Juhyun Kim^109^, Pranav Kulkarni^109^, Samuel M Lushtak^109^, Ethan Mann^109^, Max Popken^109^, Connor Soohoo^109^, Kushal Tirumala^109^, Albert Tseng^109^, Vignesh Varadarajan^109^, Jagath Vytheeswaran^109^, Christopher Wang^109^, Akshay Yeluri^109^, Dominic Yurk^109^, Michael Zhang^109^, Alexander Zlokapa^109^, Robert Pagano^110^, Chandini Jain^111^, Vishal Tomar^111^, Lam Ho^112^, Huong Huynh^113,114^, Quoc Tran^113,115^, Velma K Lopez^116^, Jo W Walker^116^, Rachel B Slayton^116^, Michael A Johansson^116^, Matthew Biggerstaff^116^, Nicholas G Reich^1^

^1^University of Massachusetts Amherst

^2^Chair of Econometrics and Statistics, Karlsruhe Institute of Technology

^3^Computational Statistics Group, Heidelberg Institute for Theoretical Studies

^4^IQT Labs

^5^Iowa State University

^6^Institute of Mathematical Statistics and Actuarial Science, University of Bern

^7^Unaffiliated

^8^Walmart

^9^Wadhwani Institute of Artificial Intelligence

^10^University of Virginia

^11^Texas Advanced Computing Center

^12^University of Texas at Austin

^13^Santa Fe Institute

^14^University of Southern California

^15^US Army Engineer Research and Development Center

^16^State University of New York Upstate Medical University

^17^Syracuse University

^18^Trinity University, San Antonio

^19^University of Michigan - Ann Arbor

^20^University of Washington

^21^University of California, San Diego

^22^Northeastern University

^23^University of California at Santa Barbara

^24^University of California, Merced

^25^Jilin University

^26^University of Science and Technology of China

^27^University of California, Los Angeles

^28^University of Chicago

^29^University of Nebraska Omaha

^30^National Cancer Institute (NCI), NIH

^31^University of Central Florida

^32^University of Arizona

^33^Texas Tech University

^34^Construx

^35^Signature Science, LLC

^36^Swiss Data Science Center, EPFL & ETHZ

^37^Institute of Global Health, Faculty of Medicine, University of Geneva

^38^Center for Intelligent Systems, EPFL

^39^Rensselaer Polytechnic Institute

^40^Arizona State University

^41^Brown University

^42^Predictive Science, Inc

^43^Imperial College, London

^44^University of Dallas

^45^Oliver Wyman

^46^University of Notre Dame

^47^Masaryk University

^48^Microsoft

^49^Laboratory for the Modeling of Biological and Socio-technical Systems, Northeastern University

^50^ISI Foundation

^51^Operations Research Center, Massachusetts Institute of Technology

^52^Sloan School of Management, Massachusetts Institute of Technology

^53^Sloan School of Management and Operations Research Center, Massachusetts Institute of Technology

^54^Leonard N Stern School of Business, New York University

^55^Institute for Data, Systems, and Society, Massachusetts Institute of Technology

^56^Laboratory for Computational Physiology, Massachusetts Institute of Technology

^57^River Hill High School

^58^SAS Institute Inc

^59^Los Alamos National Laboratory

^60^TRIUMF

^61^University of Victoria

^62^Johns Hopkins University Applied Physics Lab

^63^ School of Engineering and Applied Sciences, Harvard University

^64^Google Research

^65^Department of Epidemiology, UNC Gillings School of Public Health, University of North Carolina at Chapel Hill

^66^Harvard TH Chan School of Public Health

^67^Johns Hopkins Bloomberg School of Public Health

^68^Johns Hopkins University

^69^University of Utah

^70^École Polytechnique Fédérale de Lausanne

^71^Department of Epidemiology, University of North Carolina Gillings School of Global Public Health

^72^Indiana University–Purdue University Indianapolis

^73^University of Illinois at Urbana-Champaign

^74^Analytics Center of Excellence, IQVIA

^75^Georgia Institute of Technology

^76^IQVIA

^77^Amplitude

^78^Clemson University

^79^College of William & Mary

^80^Institute of Business Forecasting

^81^Imperial College London

^82^School of Life Sciences, University of Sussex

^83^IEM, Inc.

^84^The Hong Kong University of Science and Technology

^85^University of Iowa

^86^Virginia Tech

^87^Metron, Inc.

^88^Georgia Insitute of Technology

^89^Google Cloud

^90^Harvard University

^91^Federal Reserve Bank of San Francisco

^92^Food and Drug Administration, Center for Biologics Evaluation and Research

^93^NIH

^94^Facebook AI Research

^95^Facebook

^96^London School of Hygiene & Tropical Medicine

^97^The University of Texas at Austin

^98^Columbia University

^99^Massachusetts General Hospital

^100^Emory University Medical School

^101^Massachusetts General Hospital, Harvard Medical School

^102^Value Analytics Labs

^103^Boston University School of Medicine

^104^UNC Chapel Hill

^105^Carnegie Mellon University

^106^University of British Columbia

^107^Stanford University

^108^University of Georgia

^109^California Institute of Technology

^110^No affiliation

^111^Auquan

^112^Dalhousie University

^113^AIpert

^114^Virtual Power System

^115^Walmart Inc.

^116^Centers for Disease Control and Prevention

## Supplemental Information

**Supplement 1:** Optional fields in each metadata file:

1. **institution_affil** - University or company names, if relevant.
2. **team_funding** - Like an acknowledgement in a manuscript, you can acknowledge funding here.
3. **repo_url** - A github repository url or something similar.
4. **twitter_handles** - one or more twitter handles (without the @) separated by commas.
5. **data_inputs** - A description of the data sources used to inform the model and the truth data targeted by model forecasts. Common data sources are NYTimes, JHU CSSE, COVIDTracking, Google mobility, HHS hospitalization etc. An example description could be “cases forecasts use NYTimes data and target JHU CSSE truth data, hospitalization forecasts use and target HHS hospitalization data”
6. **citation** - a url (doi link preferred) to an extended description of your model, e.g. blog post, website, preprint, or peer-reviewed manuscript.
7. **methods_long** - An extended description of the methods used in the model. If the model is modified, this field can be used to provide the date of the modification and a description of the change.

**Supplement 2:** Validations for pull requests and metadata files submitted to the forecast hub

Each time an update (pull request) from a forecast team is submitted to the Hub, a set of validation rules that enforce the metadata requirements as outlined in the Data Records section are applied to the contents of the update. Any file in an update that fails to conform to the rules will cause the entire update to fail.

### Forecasts

Each forecast file is subject to the validation rules documented at: https://github.com/reichlab/covid19-forecast-hub/wiki/Forecast-Checks.

### Miscellaneous

Additionally, each team must have their files under a folder named consistently with their *model_abbr*, and they must only have one *primary* model.

